# Development and validation of MedDRA Tagger: a tool for extraction and structuring medical information from clinical notes

**DOI:** 10.1101/2022.12.14.22283470

**Authors:** Marie Humbert-Droz, Jessica Corley, Suzanne Tamang, Olivier Gevaert

## Abstract

Rapid and automated extraction of clinical information from patients’ notes is a desirable though difficult task. Natural language processing (NLP) and machine learning have great potential to automate and accelerate such applications, but developing such models can require a large amount of labeled clinical text, which can be a slow and laborious process. To address this gap, we propose the MedDRA tagger, a fast annotation tool that makes use of industrial level libraries such as spaCy, biomedical ontologies and weak supervision to annotate and extract clinical concepts at scale. The tool can be used to annotate clinical text and obtain labels for training machine learning models and further refine the clinical concept extraction performance, or to extract clinical concepts for observational study purposes. To demonstrate the usability and versatility of our tool, we present three different use cases: we use the tagger to determine patients with a primary brain cancer diagnosis, we show evidence of rising mental health symptoms at the population level and our last use case shows the evolution of COVID-19 symptomatology throughout three waves between February 2020 and October 2021. The validation of our tool showed good performance on both specific annotations from our development set (F1 score 0.81) and open source annotated data set (F1 score 0.79). We successfully demonstrate the versatility of our pipeline with three different use cases. Finally, we note that the modular nature of our tool allows for a straightforward adaptation to another biomedical ontology. We also show that our tool is independent of EHR system, and as such generalizable.

## INTRODUCTION

Rapid and automated extraction of clinical information from patients’ notes is a desirable though difficult task. Software and machine learning models that perform extraction of clinical concepts have been developed within the research community [1–4]. Although many are open source, they have be difficult to apply, due to the complexity of installation and require customization to a new corpus. Information such as signs and symptoms, disease severity or patient-reported outcomes are of particular interest as they can be subsequently used for phenotypic classification, clinical diagnosis, or clinical decision support [5–7].

Natural language processing (NLP) and machine learning have great potential to automate and accelerate such applications [8–11]. Developing such models requires large amount of labeled clinical text, which can be a particularly challenging task [10,12,13]. Shared resources for clinical NLP have been made available to the research community through initiatives such as MIMIC [14], National NLP Clinical Challenges (n2c2), ShARe/CLEF and SemEval. These resources help tremendously the development of machine learning systems for healthcare, though remain limited, especially when considering generalizability of models through multiple institutions.

To address this gap, we propose the MedDRA tagger, a fast annotation tool that makes use of industrial level libraries like spaCy, biomedical ontologies and weak supervision to annotate and extract clinical concepts at scale. The tool can be used to annotate clinical text and obtain labels for training machine learning models and further refine the clinical concept extraction performance, or to extract clinical concepts for observational study purposes. Our tool uses the Medical Dictionary for Regulatory Activities (MedDRA) [15] terminology as basis for annotation. MedDRA has been developed to encode and describe adverse drug events (ADE) and as such it contains an extensive description and hierarchy of signs and symptoms. It has been used to develop automatic ADE detection systems with varying levels of performance [16–21]. The simple organization of terms around system organ classes and a wide variety of lower-level terms makes MedDRA a great candidate to extract such information from clinical text.

Here, we present the MedDRA tagger and validate our tool in two ways: first, we evaluate our system on an external open-source dataset of linguistic annotations from a shared NLP task. These annotations are considered gold labels. Since the notes from this dataset come from MIMIC III, this step also assesses the generalizability of our tool to a different institution’s EMR. Second, we evaluate the performance of our system on set of randomly selected notes for three chosen psychiatric symptoms annotated by a subject matter expert. This second step allows for an extension of our validation to psychiatric disorders mentions, that are less present in the open-source dataset. We also compared the performance of our tool with the performance of cTAKES on the same validation datasets, as industry reference

Finally, to demonstrate the usability and versatility of our tool, we present three different applications: we use the MedDRA tagger to determine patients with a primary brain cancer diagnosis, we assess the prevalence of mental health symptoms at the population level and our last use case shows the evolution of COVID-19 symptomatology throughout three waves between February 2020 and October 2021. Our three applications demonstrate the versatility of MedDRA tagger with successful application in widely different areas, namely brain cancer, mental health and symptom surveillance. We were able to extract clinical concepts rapidly from millions of notes without complicated installation setup. MedDRA tagger has been able to successfully process notes from different sources without EHR-specific preprocessing or fine-tuning, thus demonstrating its versatility and ease of use.

## MATERIALS AND METHODS

### Dataset description

This study was approved by the IRB under protocol 50033: “Machine Learning of Electronic Medical Records for Precision Medicine”. The main data source that was used to conduct our study is the Stanford STARR-OMOP dataset. STARR-OMOP is Stanford Electronic Health Record data from its two Hospitals in a Observational Medical Outcomes Partnership (OMOP) Common Data Model (CDM), with linked de-identified free-text patient notes, referred to as STARR-OMOP from now on. We also obtained notes from the Stanford Cancer Institute Research Database (SCIRDB) for the brain tumor use case. Finally, we used an open-source data set for part of our validation from the ShARe/CLEF 2014 challenge [22,23]

We used two datasets for validation. First, the test set from the ShARe 2014 task 2 with gold standard annotation for disorders mentions (referred to as ShARe dataset throughout this manuscript). It contains 133 notes sampled from MIMIC-II [24]. Second, we randomly sampled 525 mentions of selected psychiatric disorders symptoms from 341 notes spanning 327 patients from the STARR OMOP dataset.

For the first use case, we used a cohort of 1,336 patients with a brain tumor diagnosis and at least one pathology report. Using the clinical data from SCIRDB, we selected only reports within approximately 60 days of diagnosis, leading to 718 patients and 1,309 reports to process with the MedDRA tagger. For the second use case, we used all the notes for all patients for the calendar year 2020 from the STARR-OMOP dataset for the analysis. To establish a baseline, we used an average of the 3 years prior (2017 – 2019). The analysis data contains 4,550,255 notes from 575,199 patients and the baseline data contains an average of 4,125,779 notes for 600,209 patients per year (Table 1). Finally, for our third use case, we selected all patients with at least one encounter with a COVID-19 diagnosis from the snapshot of our database on November 4, 2021, yielding 31,047 patients with 563,299 notes.

**Table 1:**
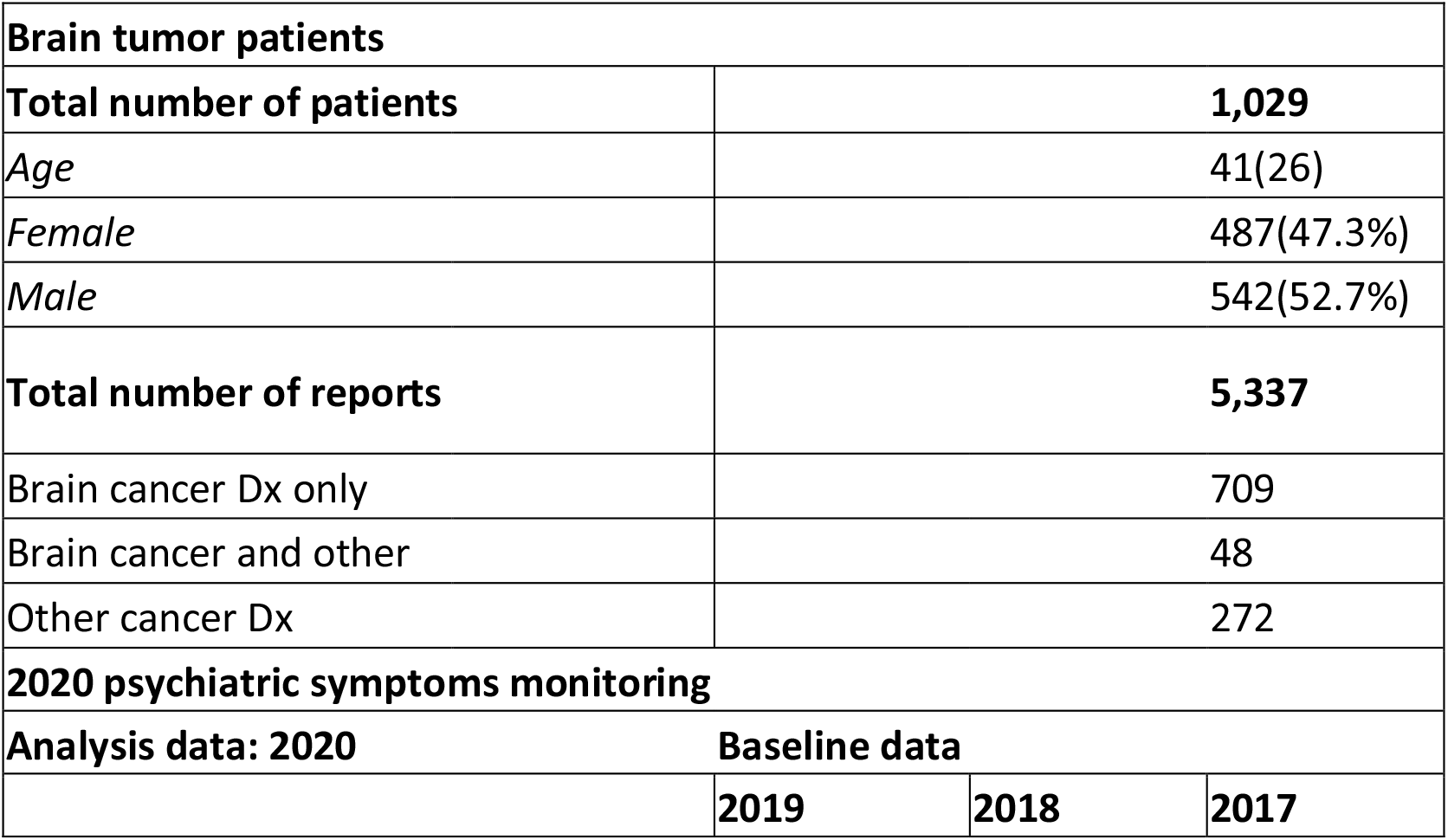
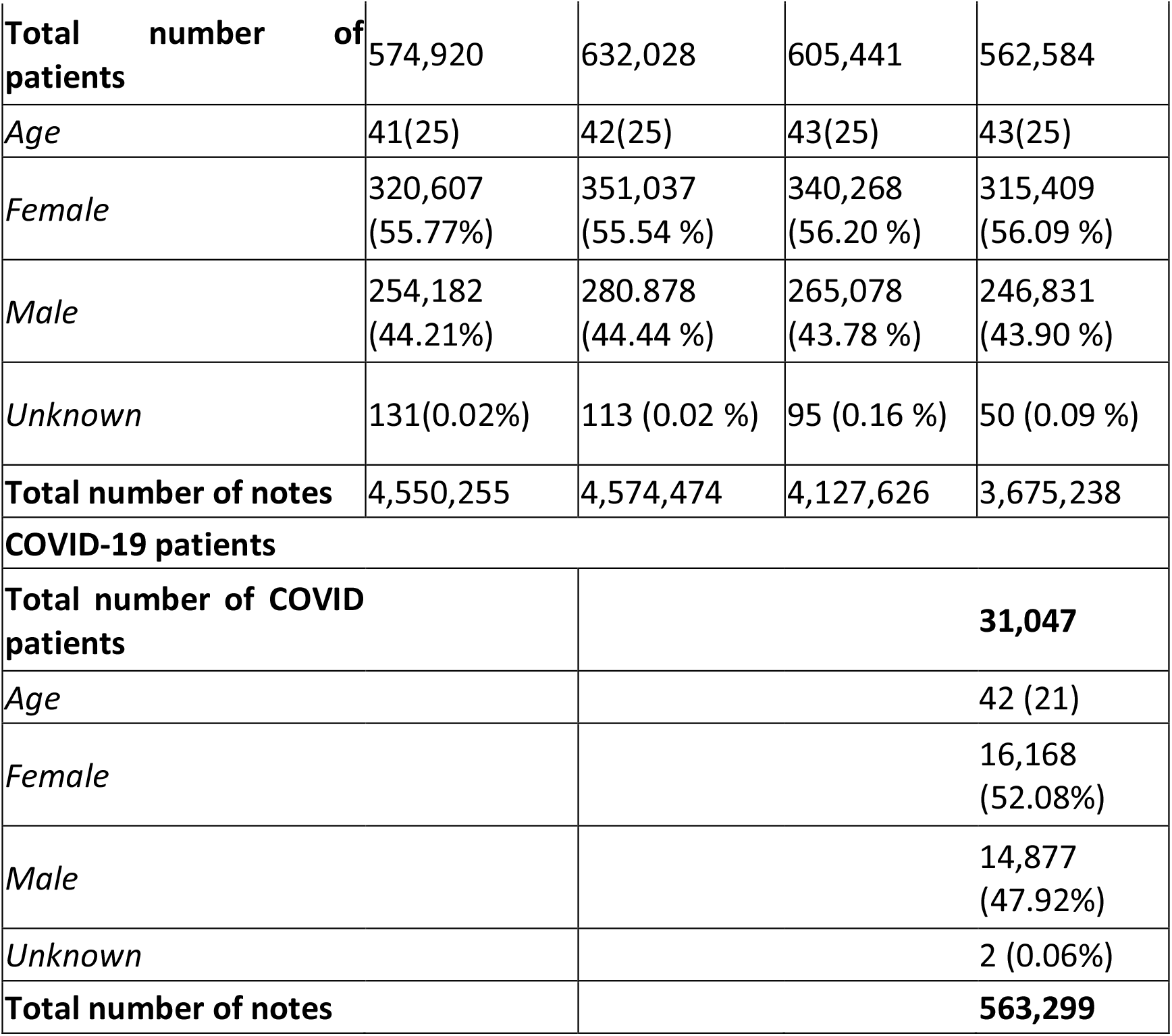
Datasets description for the three use cases.

| Brain tumor patients |  |  |  |
| --- | --- | --- | --- |
| Total number of patients | 1,029 |  |  |
| Age | 41(26) |  |  |
| Female | 487(47.3%) |  |  |
| Male | 542(52.7%) |  |  |
| Total number of reports | 5,337 |  |  |
| Brain cancer Dx only | 709 |  |  |
| Brain cancer and other | 48 |  |  |
| Other cancer Dx | 272 |  |  |
| 2020 psychiatric symptoms monitoring |  |  |  |
| Analysis data: 2020 |  | Baseline data |  |
|  | 2019 | 2018 | 2017 |

|  |  |  |  |  |
| --- | --- | --- | --- | --- |
| **Total number of patients** | 574,920 | 632,028 | 605,441 | 562,584 |
| *Age* | 41(25) | 42(25) | 43(25) | 43(25) |
| *Female* | 320,607<br>(55.77%) | 351,037<br>(55.54 %) | 340,268<br>(56.20 %) | 315,409<br>(56.09 %) |
| *Male* | 254,182<br>(44.21%) | 280.878<br>(44.44 %) | 265,078<br>(43.78 %) | 246,831<br>(43.90 %) |
| *Unknown* | 131(0.02%) | 113 (0.02 %) | 95 (0.16 %) | 50 (0.09 %) |
| **Total number of notes** | 4,550,255 | 4,574,474 | 4,127,626 | 3,675,238 |
| **COVID-19 patients** |  |  |  |  |
| **Total number of COVID patients** | **31,047** |  |  |  |
| *Age* | 42 (21) |  |  |  |
| *Female* | 16,168<br>(52.08%) |  |  |  |
| *Male* | 14,877<br>(47.92%) |  |  |  |
| *Unknown* | 2 (0.06%) |  |  |  |
| **Total number of notes** | **563,299** |  |  |  |

### Pipeline description

To extract clinical information from patient notes, we have designed a hybrid tool, taking advantage of existing NLP libraries of industrial quality and medical ontologies to annotate all clinical concepts within MedDRA [15]. A selected number of modifiers (polarity, experiencer and temporality) is determined through a weak supervision approach [25].

The MedDRA tagger pipeline is composed of three modules: the annotation module, the weak supervision labeling module, and the hierarchy mapping module (Figure 1).

**Figure 1.**
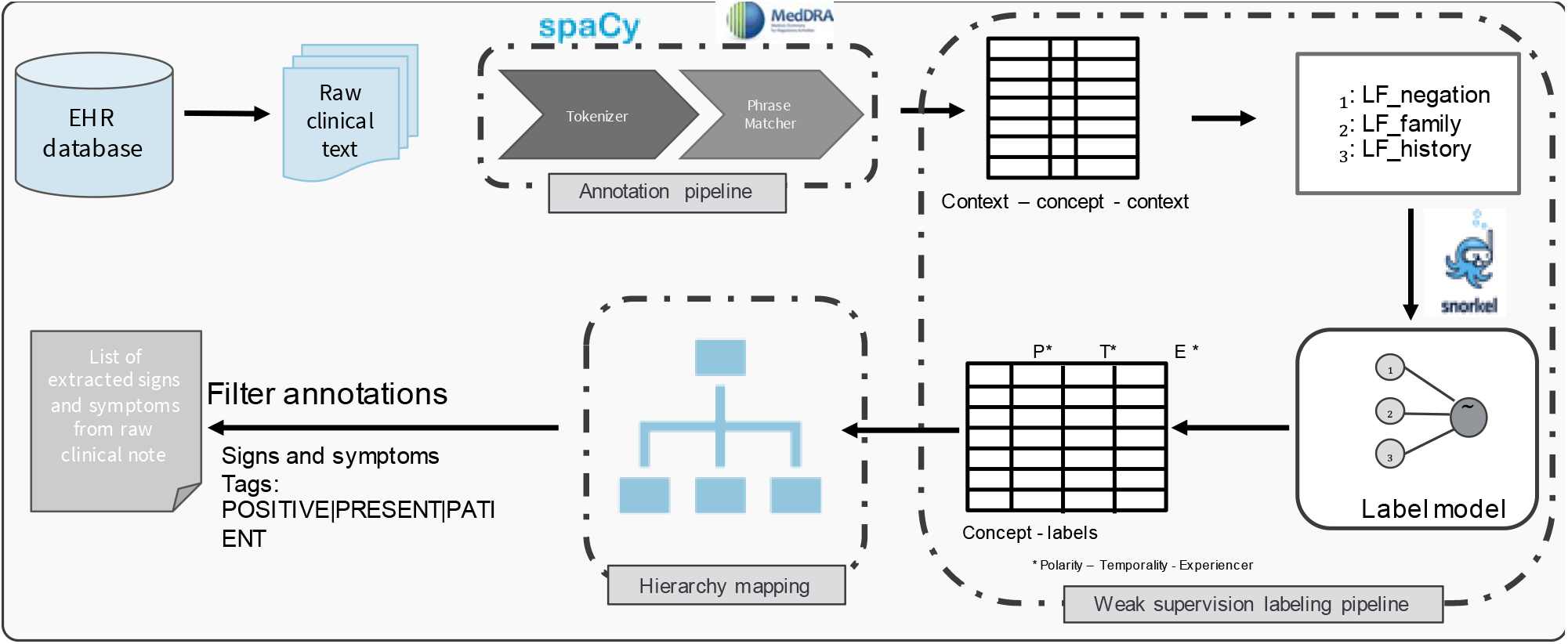
MedDRA Tagger pipeline, containing three modules: the annotation pipeline, the weak labeling pipeline, and the hierarchy module.

Annotation module: to annotate all clinical concepts efficiently, we are taking advantage of the fast implementation of core NLP functions of spacy using a ScispaCy language model [26], as well as building a custom rule-based string matching tool. We use the MedDRA terminology and match the string to the lower-case attribute of the text tokens. Each term matched by the tool is assigned its MedDRA ID number as label for further analysis.

Weak supervision module: each concept is extracted with a context window of *n* tokens before and after the concept to help determine the assertion status. A set of rule-based functions is written and applied to each extracted term and its context window using the efficiency of the Snorkel package[25]. Each term is labeled for polarity (PRESENT/ABSENT), experiencer (PATIENT/FAMILY) and temporality (PRESENT/HISTORICAL). For this project, we used previously implemented labeling functions [27] based on pattern recognition applied to a 20 tokens context window (10 tokens before and 10 tokens after the target term) to determine the negation, temporality and experiencer of the target symptom. We used the publicly available “CL-inical EVE-nt R-ecognizer” (CLEVER) base terminology [28] to match our context window with negative and hypothetical expressions (NEGEX), historical expressions (HX) and family mentions (FAM). If a mention is matched within the context window of a given term, it is labeled accordingly: ABSENT if NEGEX is matched, HISTORY if HX is matched and FAMILY if FAM is matched. Target symptoms that were positive, experienced by the patient and not part of the past medical history were labeled positive. Occurrences deviating from this pattern were labeled negative.

Finally, each term is mapped to each level of the MedDRA hierarchy using the hierarchy mapping module to ease further analysis.

### Description of MedDRA

The Medical Dictionary for Regulatory Activities (MedDRA) is a medical terminology designed for the documentation and safety monitoring of adverse drugs events. The terminology contains an extensive description and hierarchy of signs and symptoms, classified by system organ class, which makes it a well-suited terminology for the extraction of clinical concepts from patients notes. The Lowest Level Terms (LLT) allows for multiple descriptions of the same concept, usually reflecting how the term is reported in practice. Each LLT can be grouped to a Preferred Term (PT), which collects semantically equivalent terms under the same concept.

Related PTs are grouped together in the next level, the High-Level Terms (HLT) based on anatomy, pathology, physiology or function. HLTs map to Hight Level Group Terms (HLGT), which are grouped into 27 System Organ Classes (SOC), by etiology, manifestation site or purpose. This high level of granularity and grouping make the MedDRA terminology a well-suited tool for clinical concepts extraction with diverse applications.

### Validation of the MedDRA tagger

#### Using a publicly available gold standard dataset (ShAReCLEF Task2 2014)

To evaluate the performance of our tool, we have used the ShAReCLEF dataset from task 2 of the 2014 challenge [22,23]. More specifically, we used the test set from this task since it contains gold standards for entity recognition of disorders concepts. It consists of 433 de-identified clinical reports sampled from the MIMIC II dataset [24] that have been annotated for disorders mentions. Each mention has been normalized to its corresponding UMLS CUI from the SNOMED CT Disorder semantic group and modifiers have also been identified. The following modifiers have been considered for each mention: temporal expression, negation indicator, and subject.

After using our MedDRA tagger on the notes from the ShAReCLEF dataset, the extracted MedDRA terms were converted to their corresponding UMLS CUIs using the UMLS Metathesaurus [29]. We then selected only the terms within the Disorders semantic group for evaluation. Finally, we used the relations table to ensure each equivalent CUI was accounted for (using the following relations: [*isa, same_as, mapped_to*]). We then computed recall, precision and F1 for all concepts that were tagged POSITIVE, PATIENT and PRESENT by our tagger.

#### Using manually annotated notes from STARR-OMOP

The performance of the MedDRA pipeline is assessed at the mention level against manual review of 525 mentions from 341 notes. We evaluated for three psychiatric disorders concepts; anxiety, depression, and delirium. The mention evaluation was defined as follows: TP: mention is present in MedDRA output AND in annotation, TN: mention NOT present in MedDRA output AND NOT present in annotation, FP: mention present in MedDRA output and NOT present in annotation, and FN: mention NOT present in MedDRA output and present in annotation. Next, “Present” means a mention tagged POSITIVE, PRESENT, PATIENT and “NOT present” means any combination of label that differ from the definition of Present. The resulting precision, recall and F1 scores are computed at the mention level.

### Three diverse use cases of the MedDRA Tagger

To demonstrate the variety of potential applications of our tool, we present three applications. First, we make use of the MedDRA hierarchy to determine which patients with a brain tumor have a primary brain cancer from pathology notes. Second, we make use of the fast note processing power of our tool to tag all MedDRA concepts on all notes of STARR-OMOP dataset to highlight the sharp increase in psychiatric disorders during the first wave of the COVID-19 pandemic in spring 2020. Finally, we analyzed the terms extracted from the notes of COVID-19 patients throughout the pandemic to highlight the linguistic shift with the advancement of the crisis and the evolution of the waves of infection.

#### Use case 1: brain tumor diagnosis

We selected a cohort of 1,593 patients that were diagnosed with a brain tumor at Stanford with 5,337 pathology reports. Since some patients had multiple pathology reports over several years, we processed the ones that were written within 60 days of diagnosis. We used diagnoses from the Stanford Cancer Institute Research Database as our ground truth. After processing 1,309 pathology notes with the tagger, we identified notes with positive mentions of brain cancer. To do so, we took advantage of the hierarchy mapping module of the tagger and selected terms that belonged to the high level group term “Nervous system neoplasms malignant and unspecified NEC” or “Nervous system neoplasms benign” to assign a positive label to the patient.

#### Use case 2: monitoring of psychiatric symptoms in the Stanford hospital population during the COVID-19 pandemic

We have analyzed all notes from all Stanford patients for the years 2017-20 20. A total of 16,927,593 notes were processed with our MedDRA tagger to extract clinical concepts. Concepts labeled POSITIVE, PATIENT and PRESENT were selected for our analysis. Our analysis is threefold: First, all terms belonging to the psychiatric disorders SOC were selected using the hierarchy mapping module. We obtained the term count per patient and date and performed a seven-day moving average to plot the term counts over time. Term counts for the years 2017-2019 were averaged to establish a baseline. Second, term counts were calculated at the PT level. The difference between the 2020 counts and the baseline counts was plotted with a seven-day moving average. Finally, the top 7 extracted terms (LLT) are shown together with the top 7 psychiatric diagnoses from the coded part of EHR.

#### Use case 3: detecting common symptoms of COVID19 patients

We have selected all encounters with a diagnosis of COVID-19 and analyzed the corresponding notes using the MedDRA tagger and a TFIDF-based scoring system to highlight the most informative terms at a certain point in time. To identify important terms among patients within a certain timeframe, we used a scoring system based on term frequency-inverse document frequency (TFIDF). We modified the original definition of TFIDF in the following manner: for each term in the corpus, we compute a TFIDF, based on each month of data. For the considered month:

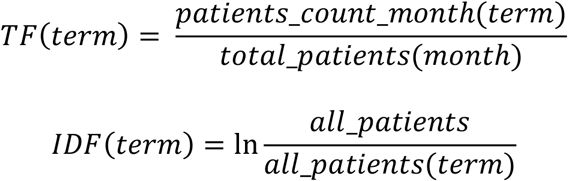

We selected the top 6 ranked terms according to their modified-TFIDF scoring for three chosen months where infection rates were high in the STARR-OMOP database; June 2020, December 2020 and August 2021 (Table 2). We then plotted the resulting score for each term throughout the time frame of our dataset (February 2020 – October 2021) to highlight trends in terms frequency according to each wave. We also added the visit counts associated to a COVID-19 diagnosis to the resulting graph to identify the different waves of infection.

**Table 2.** Application 3 - Top terms selected for TFIDF analysis

| **June 2020** | **December 2020** | **August 2021** |
| --- | --- | --- |
| Diabetes | Headache | Chest pain |
| Feverish | Home isolation | Confusion |
| Loss of taste | Immunocompromised | Diarrhea |
| Nausea | Quarantine | Home quarantine |
| Sore throat | Respiratory distress | Hypertension |
|  | Shortness of breath | Vaccination |

## RESULTS

### Validation using standard data set – ShAReCLEF task 2 2014

To evaluate the performance of our tool, we used the ShARe dataset since it was annotated for all the disorders mentions present in discharge summaries from the MIMIC II dataset. Each mention was also normalized to standard vocabulary, making the comparison with our annotations straightforward. The resulting performance was a recall of 0.86, precision of 0.73 and F1 of 0.79.

### Validation using 341 notes from STARR-OMOP dataset with expert annotation

The performance of the MedDRA tagger was then assessed against manual review of 525 mentions from 341 notes. We calculated the performance of our tool at the mention level for three psychiatric symptoms. Mentions were randomly selected from a set of 341 notes from the STARR-OMOP dataset. The resulting performance was a recall of 0.77, precision of 0.85 and F1 of 0.81 (Table 3).

**Table 3.** Performance of the MedDRA tagger on 2 validation datasets

| Concept | Number of mentions evaluated |  | Recall | Precision | F1 |
| --- | --- | --- | --- | --- | --- |
| **Anxiety** | 223 | MedDRA tagger | 0.82 | 0.92 | 0.87 |
|  |  | cTAKES | 0.92 | 0.68 | 0.78 |
| **Depression** | 132 | MedDRA tagger | 0.69 | 0.91 | 0.78 |
|  |  | cTAKES | 0.96 | 0.71 | 0.82 |
| **Delirium** | 170 | MedDRA tagger | 0.79 | 0.97 | 0.87 |
|  |  | cTAKES | 0.89 | 0.69 | 0.78 |
| **Total psychiatric mentions** | **525** | **MedDRA tagger** | **0.77** | **0.85** | **0.81** |
|  |  | **cTAKES** | **0.93** | **0.56** | **0.70** |
| **ShARe dataset** | **6,736** | **MedDRA tagger** | **0.86** | **0.73** | **0.79** |
|  |  | **cTAKES** | **1.0** | **0.18** | **0.29** |

**Table 4.** Syndromic surveillance of mental health symptoms: Patient count for top psychiatric terms from diagnosis table and NLP extraction

| CONCEPT | NB PATIENTS | SOURCE |
| --- | --- | --- |
| ANXIETY DISORDER | 22,993 | DX |
| MAJOR DEPRESSION, SINGLE EPISODE | 19,504 | DX |
| DEPRESSIVE DISORDER | 12,031 | DX |
| GENERALIZED ANXIETY DISORDER | 11,129 | DX |
| MODERATE RECURRENT MAJOR DEPRESSION | 2,491 | DX |
| PANIC DISORDER WITHOUT AGORAPHOBIA | 2,029 | DX |
| MODERATE MAJOR DEPRESSION, SINGLE EPISODE | 1,171 | DX |
| DEPRESSION | 147,305 | NLP |
| STRESS | 123,122 | NLP |
| ANXIETY DISORDER | 121,636 | NLP |
| FEELING DOWN | 87,772 | NLP |
| VIOLENCE | 47,736 | NLP |
| WORRY | 35,142 | NLP |

### Application 1: Determination of primary site for cancer patients from pathology notes

In order to automatically detect the primary cancer site for patients from their pathology notes, we processed all pathology reports from our cohort of patients with brain cancer. Using the primary site information from our clinical data base as gold standard, our tool showed a precision score of 0.95, a recall score of 0.87 and an F1 score of 0.90.

### Application 2: Syndromic surveillance: the case for mental health symptoms extraction

In order to highlight psychiatric effects of a global pandemic and make the case for inclusion of mental health symptoms in syndromic surveillance systems, we processed all the notes for the year 2020 as well as the baseline with the MedDRA tagger to extract all clinical concepts from the notes. Figure 2a shows concepts counts per patient and date with a seven day moving average for all concepts belonging to the psychiatric disorders system organ class. The mentions show a rapid increase starting in March 2020 at the beginning of the pandemic, reaching a peak in April 2020 and returning to baseline in June 2020. The breakdown of the mentions counts at the preferred term level highlights that anxiety symptoms are the driver of this sharp increase (Figure 2b). Finally, a comparison of the top seven psychiatric mentions in the text with the top seven diagnosis codes from the encounter data shows a tenfold difference in frequency, reinforcing the initial hypothesis that mental health information is better documented in clinical notes than in the coded part of EHR (Figure 2c).

**Figure 2.**
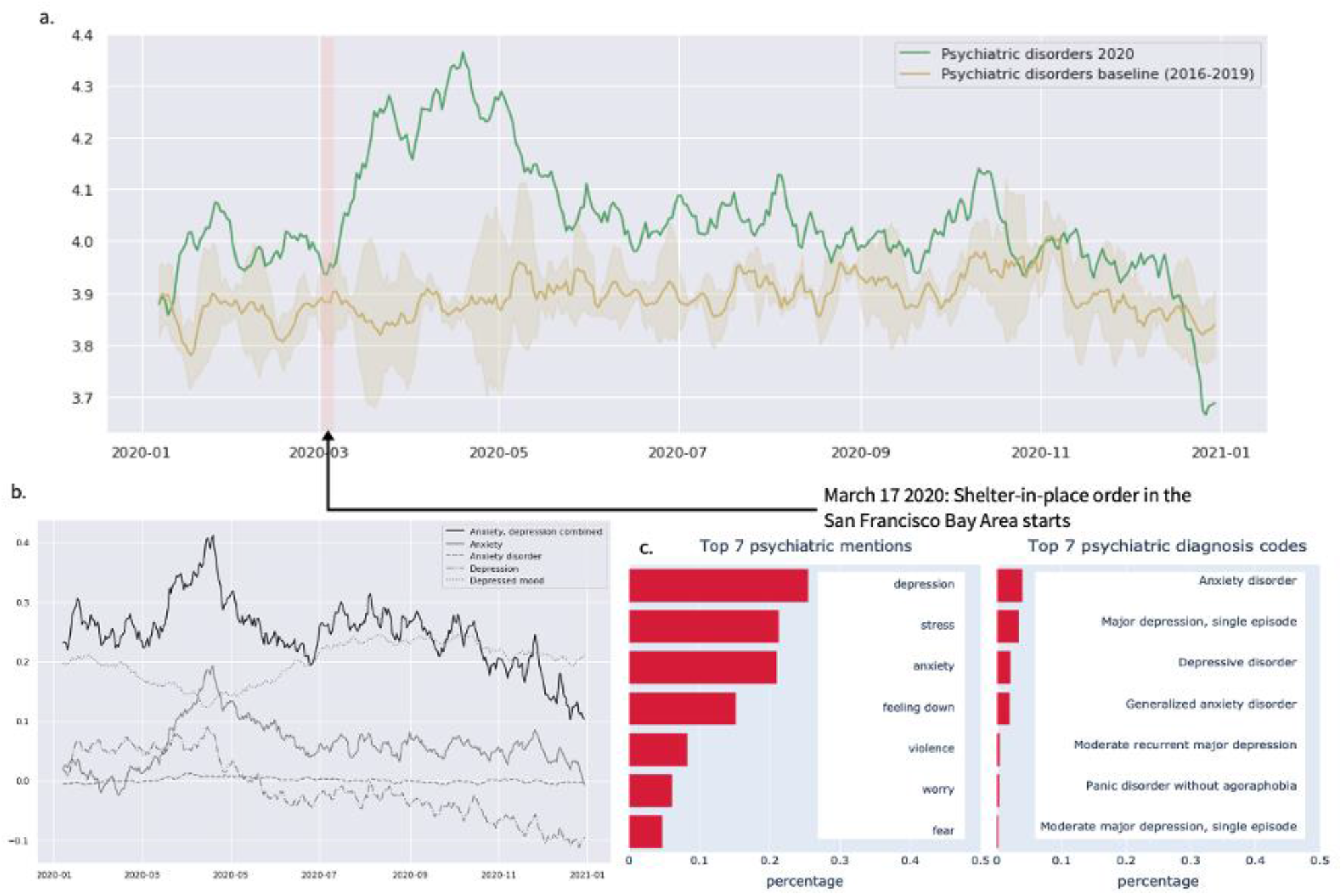
Syndromic surveillance of mental health symptoms. a) Psychiatric disorders mentions counts normalized per patient and date with a 7-days moving average. b) Difference of mentions counts at the preferred term level (2020 – baseline) for a selection of terms – Anxiety, anxiety disorder, depression, depressed mood. c) Top 7 extracted mentions and top 7 psychiatric diagnosis codes.

### Application 3: Symptomatology of COVID patients – evolution of symptoms with different waves and dominant variants of COVID

In the context of the pandemic, we also analyzed common symptoms among patients to highlight trends associated with each wave of infection. We selected six top terms identified during each wave of the COVID pandemic within Stanford’s patients between February 2020 and October 2021. Top terms for June 2020, December 2020 and August 2021 were chosen and their respective TFIDF score for each month is plotted on Figure 3. For reference, the visits with a COVID-19 diagnosis are also displayed on the figure.

**Figure 3.**
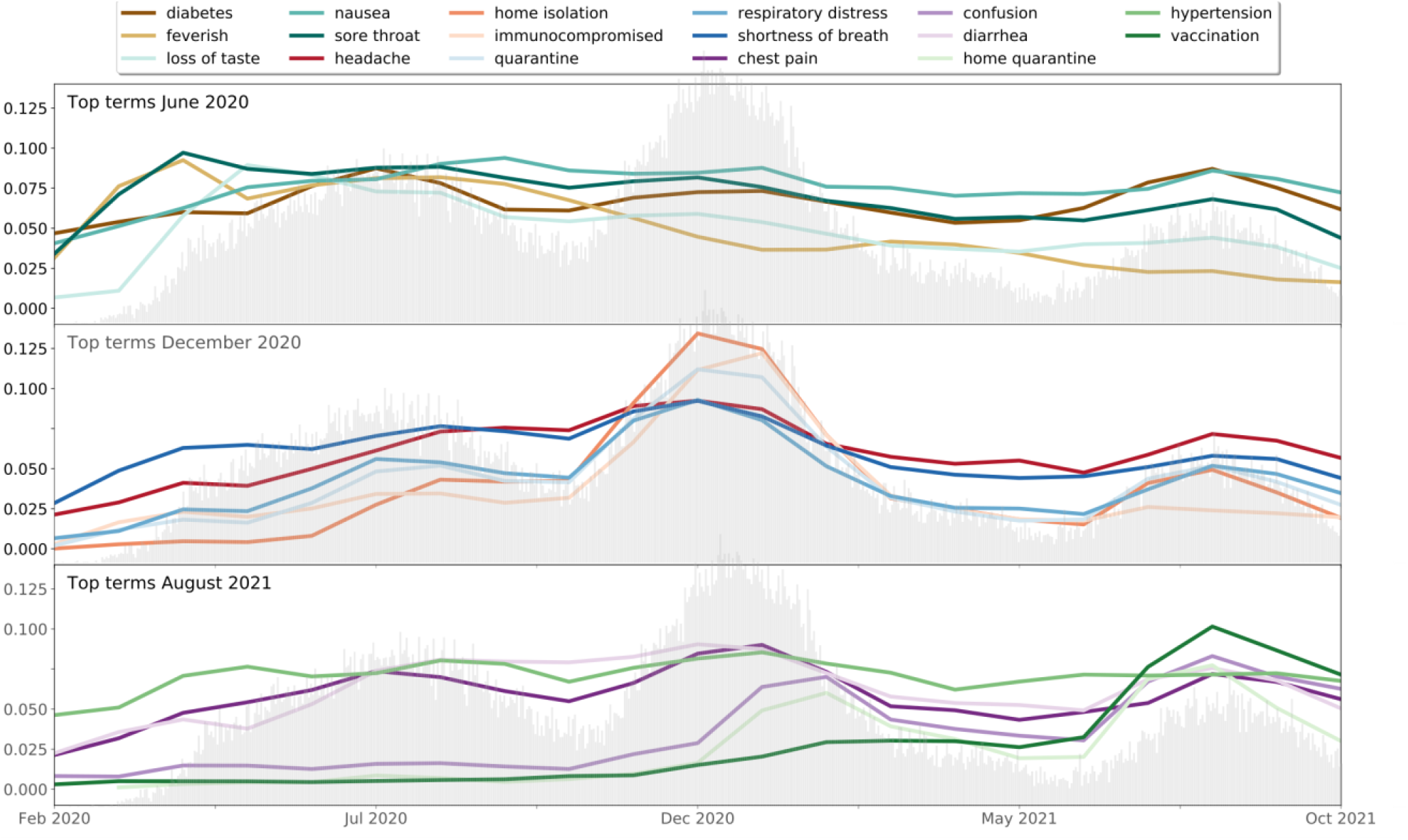
Symptomatology of COVID-19 patients: TFIDF scores of terms selected throughout the pandemic

Terms identified from the first wave of infections are mostly common symptoms that could be attributed to many infectious diseases. Their score remains consistent throughout the whole considered period, except for loss of taste, which is specific to COVID-19, that sharply rises with the first wave. The second wave shows the appearance of terms that were new at the beginning of the pandemic, but became associated with the pandemic by the end of 2020, like home isolation and quarantine. The rise in use of the immunocompromise term was also detected by the MedDRA tagger, as it has been shown that immunocompromised people are at higher risk of COVID-19 infections and severe outcomes [30–33]. Finally, terms of the third wave reflect the emergence of vaccines, with vaccination the term with the highest TFIDF score. Home quarantine is also a very important term. Finally, the third wave shows the appearance of neurological function terms like confusion, which are also prevalent among COVID-19 infections but were identified later on.

## DISCUSSION

We developed and evaluated a novel pipeline for extracting clinical concepts from medical notes leveraging the MedDRA terminology. The pipeline showed good performance on both strategies for validation. Finally, we demonstrated the usability of the tool on three different use cases.

The mention-level performance of our tool is comparable to popular software like cTAKES. However, our tool differs from cTAKES in software design, scope and functionality. There is no installation necessary beyond the required python libraries to run the scripts. By making use of industrial-level libraries, the processing of notes is fast and computationally inexpensive. For example, the processing of the 2020 data in use case 2 (>4.5 million documents) took only 50h using 32 CPUs. The output is straightforward and easy to process in downstream analytic applications. On the contrary, more complex tools, even though offering more information about entities, co-reference functionality and additional information such as relationships, can be difficult for a researcher to install, slower to process each document, and the output needs further processing to analyze the extracted mentions.

The analysis of false positives shows that some improvements could be made with the negation labeling functions. Indeed, when the concept of interest is within a list of negated conditions, as is often the case in clinical notes, the negation may be located outside of the context window. Increasing this window may improve the false positive rate.

MedDRA has been developed to facilitate sharing of regulatory information for medical products pre- and post-market. Its rich vocabulary allows for precise reporting of adverse reaction in the context of pharmacovigilance. Past work using MedDRA and NLP methods for information extraction have focused on adverse drug event (ADE) recognition. For example, Zorzi et al. developed a NLP pipeline for processing notes and linking them to MedDRA in Italian language for automatic detection of ADE [17–19]. They used an annotated dataset of 4,500 ADE from an Italian pharmacovigilance data warehouse. Their system showed a performance of 65-69% recall and 61-70% precision. Wang et al. evaluated the performance of four systems for the recognition of symptomatic adverse events and normalized them to MedDRA from patient-authored text. The best system for strict matching shoed a F1 score of 0.56 [34]. Ly et al. [20] used NLP methods and MedDRA to annotate drug product labeling. They evaluated three NLP systems for adverse events extraction and mapping to MedDRA preferred text. Precision ranged from 64%-77%, recall from 64%-83% and F1 from 67%-79%. Though the task is different, it is worth noting that our MedDRA tagger outperforms all these systems for the mention recognition.

Because of the rich vocabulary describing signs and symptoms, we believe MedDRA is well suited for tasks beyond adverse drug event recognition. Indeed, on a more general task of extracting disorders mentions in clinical notes, our tool showed great performance, with recall of 86%, precision of 73% and F1 score of 79% on a standard dataset. To our knowledge, our system is the first tool that makes use of MedDRA for such wider purpose. Furthermore, we demonstrate an extensive range of applicability and the validation using two datasets from separate institutions shows great generalizability. The tool is easy to use and does not require installation beyond the required python libraries to run the scripts. Finally, the simple and modular pipeline based on SpaCy allows for straightforward extensions to other ontologies or even multiple languages

## CONCLUSION

Throughout this paper, we present the development and validation of a simple yet efficient NLP tool for extraction of clinical concepts from text. We show the versatility of our tool with three use case examples, ranging from brain cancer to infectious diseases, demonstrating that its use is not restricted to a specific disease area. We have chosen to use MedDRA for its rich low level terms that more likely to be used in clinical practice. We note that the modular nature of our tool allows for a straightforward adaptation to another biomedical ontology. We also show that our tool is independent of EHR system, and as such generalizable.

## Data Availability

This research used data from clinical notes from the Stanford Children's Hospital, the University Healthcare Alliance and Packard Children's Health Alliance clinics. Due to the presence of private health identifiers in these notes, the raw data cannot be shared publicly.

## DATA AVAILABILITY

This research used data from clinical notes from the Stanford Children’s Hospital, the University Healthcare Alliance and Packard Children’s Health Alliance clinics. Due to the presence of PHI data in these notes, the raw data cannot be shared publicly.

## AUTHOR CONTRIBUTIONS

Concept: Marie Humbert-Droz, Olivier Gevaert. Data curation: Marie Humbert-Droz, Jessica Corley. Data Analysis, Coding: Marie Humbert-Droz. Supervision: Olivier Gevaert, Suzanne Tamang. Manuscript Writing: Marie Humbert-Droz, Jessica Corley, Suzanne Tamang, Olivier Gevaert.

## REFERENCES

1. Savova GK, Masanz JJ, Ogren P V., Zheng J, Sohn S, Kipper-Schuler KC, Chute CG. Mayo clinical Text Analysis and Knowledge Extraction System (cTAKES): Architecture, component evaluation and applications. J Am Med Informatics Assoc 2010 Sep;17(5):507–513. PMID:20819853

2. Soysal E, Wang J, Jiang M, Wu Y, Pakhomov S, Liu H, Xu H. CLAMP - a toolkit for efficiently building customized clinical natural language processing pipelines. J Am Med Informatics Assoc Oxford University Press; 2018 Mar 1;25(3):331–336. PMID:29186491

3. Aronson AR, Lang FM. An overview of MetaMap: Historical perspective and recent advances. J Am Med Informatics Assoc 2010;17(3):229–236. PMID:20442139

4. Chiang JH, Lin JW, Yang CW. Automated evaluation of electronic discharge notes to assess quality of care for cardiovascular diseases using Medical Language Extraction And Encoding System (MedLEE). J Am Med Informatics Assoc 2010;17(3):245–252. PMID:20442141

5. Koleck TA, Dreisbach C, Bourne PE, Bakken S. Natural language processing of symptoms documented in free-text narratives of electronic health records: A systematic review. J Am Med Informatics Assoc. Oxford University Press; 2019. p. 364–379. PMID:30726935

6. Forbush TB, Gundlapalli A V, Palmer MN, Shen S, South BR, Divita G, Carter M, Redd A, Butler JM, Samore M. “Sitting on pins and needles”: characterization of symptom descriptions in clinical notes”. AMIA Jt Summits Transl Sci proceedings AMIA Jt Summits Transl Sci 2013;2013:67–71. PMID:24303238

7. Adnan K, Akbar R, Khor, S. W, Ali, A. B. A. Role and challenges of unstructured big data in healthcare. Data Manag Anal Innov 2020;301–323.

8. Chapman WW, Nadkarni PM, Hirschman L, D’Avolio LW, Savova GK, Uzuner O. Overcoming barriers to NLP for clinical text: The role of shared tasks and the need for additional creative solutions. J Am Med Informatics Assoc. 2011. p. 540–543. PMID:21846785

9. Friedman C, Rindflesch TC, Corn M. Natural language processing: State of the art and prospects for significant progress, a workshop sponsored by the National Library of Medicine. J Biomed Inform 2013 Oct;46(5):765–773. PMID:23810857

10. Velupillai S, Suominen H, Liakata M, Roberts A, Shah AD, Morley K, Osborn D, Hayes J, Stewart R, Downs J, Chapman W, Dutta R. Using clinical Natural Language Processing for health outcomes research: Overview and actionable suggestions for future advances. J Biomed Inform. Academic Press Inc.; 2018. p. 11–19. PMID:30368002

11. Patel R, Tanwani S. Application of Machine Learning Techniques in Clinical Information Extraction. Springer Int Publ Cham 2019;145–165.

12. Xiao C, Choi E, Sun J. Opportunities and challenges in developing deep learning models using electronic health records data: A systematic review. J Am Med Informatics Assoc. Oxford University Press; 2018. p. 1419–1428. PMID:29893864

13. Spasic I, Nenadic G. Clinical text data in machine learning: Systematic review. JMIR Med Informatics. JMIR Publications Inc.; 2020. [doi: 10.2196/17984]

14. Johnson AEW, Pollard TJ, Shen L, Lehman LWH, Feng M, Ghassemi M, Moody B, Szolovits P, Anthony Celi L, Mark RG. MIMIC-III, a freely accessible critical care database. Sci Data 2016;3:1–9. PMID:27219127

15. International Conference on Harmonisation of Technical Requirements for Registration of Pharmaceuticals for Human Use (ICH), MedDRA Data Retrieval and Presentation: Points to consider. 2016.

16. Bayer S, Clark C, Dang O, Aberdeen J, Brajovic S, Swank K, Hirschman L, Ball R. ADE Eval: An Evaluation of Text Processing Systems for Adverse Event Extraction from Drug Labels for Pharmacovigilance. Drug Saf Adis; 2021 Jan 1;44(1):83–94. PMID:33006728

17. Zorzi M, Combi C, Lora R, Pagliarini M, Moretti U. Automagically encoding adverse drug reactions in MedDRA. Proc - 2015 IEEE Int Conf Healthc Informatics, ICHI 2015 Institute of Electrical and Electronics Engineers Inc.; 2015. p. 90–99. [doi: 10.1109/ICHI.2015.18]

18. Combi C, Zorzi M, Pozzani G, Moretti U, Arzenton E. From narrative descriptions to MedDRA: automagically encoding adverse drug reactions. J Biomed Inform Academic Press Inc.; 2018 Aug 1;84:184–199. PMID:29981491

19. Zorzi M, Pozzani G, Combi C, Moretti U. Mapping free text into MedDRA by Natural Language Processing: A modular approach in designing and evaluating software extensions. ACM-BCB 2017 - Proc 8th ACM Int Conf Bioinformatics, Comput Biol Heal Informatics Association for Computing Machinery, Inc; 2017. p. 27–35. [doi: 10.1145/3107411.3107431]

20. Ly T, Pamer C, Dang O, Brajovic S, Haider S, Botsis T, Milward D, Winter A, Lu S, Ball R. Evaluation of Natural Language Processing (NLP) systems to annotate drug product labeling with MedDRA terminology. J Biomed Inform. Academic Press Inc.; 2018. p. 73–86. PMID:29860093

21. Friedman C. Discovering Novel Adverse Drug Events Using Natural Language Processing and Mining of the Electronic Health Record. LNAI. 2009.

22. Mowery D. ShAReCLEF eHealth Evaluation Lab 2014 (Task 2): Disorder Attributes in Clinical Reports (version 1.0). PhysioNet. 2013. [doi: 10.13026/0zgk-9j94]

23. Mowery DL, Velupillai S, South BR, Christensen L, Martinez D, Kelly L, Goeuriot L, Elhadad N, Pradhan S, Savova G, Chapman WW. Task 2: ShARe/CLEF eHealth evaluation lab 2014. CEUR Workshop Proc 2014;1180:31–42.

24. Saeed M, Villarroel M, Reisner AT, Clifford G, Lehman LW, Moody G, Heldt T, Kyaw TH, Moody B, Mark RG. Multiparameter intelligent monitoring in intensive care II: A public-access intensive care unit database. Crit Care Med Lippincott Williams and Wilkins; 2011. p. 952–960. PMID:21283005

25. Ratner A, Bach SH, Ehrenberg H, Fries J, Wu S, Ré C. Snorkel: rapid training data creation with weak supervision. VLDB J Springer; 2020. p. 709–730. [doi: 10.1007/s00778-019-00552-1]

26. Neumann M, King D, Beltagy I, Ammar W. ScispaCy: Fast and Robust Models for Biomedical Natural Language Processing. 2019.

27. Humbert-Droz M, Mukherjee P, Gevaert O. Strategies to Address the Lack of Labeled Data for Supervised Machine Learning Training With Electronic Health Records: Case Study for the Extraction of Symptoms From Clinical Notes. JMIR Med Informatics 2022;10(3):e32903. [doi: 10.2196/32903]

28. Tamang S. CLEVER base terrminology.

29. Bodenreider O. The Unified Medical Language System (UMLS): Integrating biomedical terminology. Nucleic Acids Res 2004;32(DATABASE ISS.):267–270. PMID:14681409

30. Dai M, Liu D, Liu M, Zhou F, Li G, Chen Z, Zhang Z, You H, Wu M, Zheng Q, Xiong Y, Xiong H, Wang C, Chen C, Xiong F, Zhang Y, Peng Y, Ge S, Zhen B, Yu T, Wang L, Wang H, Liu Y, Chen Y, Mei J, Gao X, Li Z, Gan L, He C, Li Z, Shi Y, Qi Y, Yang J, Tenen DG, Chai L, Mucci LA, Santillana M, Cai H. Patients with cancer appear more vulnerable to SARS-CoV-2: A multicenter study during the COVID-19 outbreak. Cancer Discov American Association for Cancer Research Inc.; 2020 Jun 1;10(6):783. PMID:32345594

31. Tian J, Yuan X, Xiao J, Zhong Q, Yang C, Liu B, Cai Y, Lu Z, Wang J, Wang Y, Liu S, Cheng B, Wang J, Zhang M, Wang L, Niu S, Yao Z, Deng X, Zhou F, Wei W, Li Q, Chen X, Chen W, Yang Q, Wu S, Fan J, Shu B, Hu Z, Wang S, Yang XP, Liu W, Miao X, Wang Z. Clinical characteristics and risk factors associated with COVID-19 disease severity in patients with cancer in Wuhan, China: a multicentre, retrospective, cohort study. Lancet Oncol Lancet Publishing Group; 2020 Jul 1;21(7):893–903. PMID:32479790

32. Baek MS, Lee MT, Kim WY, Choi JC, Jung SY. COVID-19-related outcomes in immunocompromised patients: A nationwide study in Korea. PLoS One Public Library of Science; 2021 Oct 1;16(10 October). PMID:34597325

33. Li X, Xu S, Yu M, Wang K, Tao Y, Zhou Y, Shi J, Zhou M, Wu B, Yang Z, Zhang C, Yue J, Zhang Z, Renz H, Liu X, Xie J, Xie M, Zhao J. Risk factors for severity and mortality in adult COVID-19 inpatients in Wuhan. J Allergy Clin Immunol Mosby Inc.; 2020 Jul 1;146(1):110–118. PMID:32294485

34. Wang Y, Gots D, Basch EM, Chung AE. An Evaluation of Clinical Natural Language Processing Systems to Extract Symptomatic Adverse Events from Patient-Authored Free-Text Narratives. Proc 2021 AMIA Informatics Summit Institute of Electrical and Electronics Engineers Inc.; 2021. [doi: 10.1109/ICMLA.2019.00297]

